# Towards automated fetal brain biometry reporting for 3-dimensional T2-weighted 0.55-3T magnetic resonance imaging at 20-40 weeks gestational age range

**DOI:** 10.1101/2025.02.06.25321808

**Authors:** Aysha Luis, Alena Uus, Jacqueline Matthew, Sophie Arulkumaran, Alexia Egloff Collado, Vanessa Kyriakopoulou, Sara Neves Silva, Jordina Aviles Verdera, Megan Hall, Simi Bansal, Sarah McElroy, Kathleen Colford, Joseph V. Hajnal, Jana Hutter, Lisa Story, Mary Rutherford

## Abstract

**Background:** The detailed assessment of fetal brain maturation and development involves morphological evaluation, gyration analysis, and reliable biometric measurements. Manual measurements on conventional 2-D magnetic resonance imaging (MRI) are affected by fetal motion and there is no clear consensus regarding definitions for brain biometric parameters and anatomical landmark placements, making consistent reference plane and slice selection challenging. Automated biometry with 3-D slice-to-volume reconstruction (SVR) has the potential to improve the reliability of derived measurements, allowing precise quantification of fetal brain development. Previous published works have primarily focused on the technical feasibility of automated fetal brain biometry methods for T2-weighted (T2W) MRI. However, none have proposed solutions for automating the reporting of biometry results, which could enhance clinical utility and support real-time integration into routine clinical workflows. Furthermore, there is no consensus on a universal fetal biometry protocol for 3D fetal MRI.

**Aim:** To formalise biometry protocol for 3-D SVR T2W fetal brain MRI and to develop and validate a fully automated biometry reporting pipeline.

**Materials and methods:** Automated extraction of 13 routinely reported linear fetal biometry measurements using deep learning localisation of anatomical landmarks in 3-D reconstructed T2W brain images and presentation of the results in .html report with centile calculation. The automated biometry method was tested on 90 retrospective cases and the fully automated, end-to-end biometry reporting pipeline was prospectively evaluated on 111 cases across a wide range of gestational ages, field strengths and scanning parameters. We also generated normal centile ranges for 19 - 40 weaks GA range from 406 normal control datasets.

**Results:** The retrospective evaluation showed excellent to good localisation for the majority of cases, with the maximum absolute difference between automated vs. manual measurement within 1-3mm range. For the prospective evaluation, more than 98% of all landmark placement were graded as acceptable for interpretation and measurements. The processing time of the pipeline was less than five minutes per case, with the measurements and centiles available at the time of reporting. Inspection of the automated landmark placement and computed biometrics took 1-3 minutes per case. The generated normative growth charts demonstrate high correlation with the trends in the previously reported works.

**Conclusion:** Our approach is the first to develop a pipeline which integrates automated fetal brain biometric measurements, centile calculations and normative growth charts into the clinical workflow, allowing fully automated biometry reporting for T2-weighted motion-corrected SVR MRI.

## Introduction

The role of fetal magnetic resonance imaging (MRI) has grown exponentially in recent years, and is currently used as a problem solving, prognostication and treatment planning tool [1, 2]in an ever-increasing range of applications, including fetal brain development [3, 4], fetal body anomalies[5, 6], congenital heart disease[7], and placental pathologies[8].

Accurate and detailed assessment of fetal brain maturation and development involves morphological evaluation, gyration analysis, and reliable biometric measurements. This enables early detection and characterization of several central nervous system (CNS) anomalies, allowing comprehensive antenatal and postnatal evaluation and provides a clearer understanding of the severity and impact of detected abnormalities, which results in increased parental choices and improved neonatal and childhood outcomes [9].

In current clinical practice, biometric MRI measurements are usually performed manually on 2-dimensional (2D) slices, based on the widely available methodology and reference biometric charts [10, 11] There are however inherent challenges of acquiring accurate measurements on 2D slices. Fetal and maternal motion can result in suboptimal acquisition planes, as well as inter-slice, and inter-plane motion, which can lead to false positive or false-negative results and contribute to the uncertainty in the obtained measurements. These technical challenges are particularly pronounced at early gestational ages (GA), where increased fetal movement and small brain structures make them especially susceptible to measurement error. /Although additional reference biometric charts have since been published for posterior fossa [12, 13], cavum septum pellucidum [14] and brain biometry for early GA (20-24 weeks)[15], they too rely on measurements from 2D slices.

3D slice-to-volume (SVR) reconstruction [16] of the fetal brain addresses many of the technical challenges by enabling full volumetric assessment, better visualization of fetal brain anatomy and reliable biometric measurements that are concordant with those obtained from true orthogonal 2D acquisition planes [17–19]. However, clinical adoption of 3D SVR is not as widespread despite the availability of normative reference charts and centile calculator [17].

Currently, there remains significant variation in practice across different centers with no clear consensus regarding several aspects of fetal brain biometry. These include the definition of anatomical landmarks for each measurement, selection of reference plane as well as slice, determining which structures and parameters to be measured in order to assess normal growth and diagnose specific pathologies, and deciding whether inner or outer anatomical boundaries should be used. The impact of partial volume effects and asymmetry of brain structures remain also inadequately addressed. Furthermore, there is a well-known and expected degree of variance in manual measurements, further complicating the consistency and accuracy of these assessments.

The lack of universally accepted biometry measurement definitions and methodology across different GAs and field strengths highlights the need for detailed, systematic, and standardized guidelines that can serve as a baseline. Indeed, a systematic review focused on posterior fossa measurements in fetal MRI [20] identified 62 distinct 2D biometric measurements, many of which assessed similar features using slightly different methods. Another systematic review [21] highlighted that the existing MRI reference ranges for fetal brain biometry have low-to-moderate methodological quality, emphasizing the need for a more robust, standardized approach. Similar findings were reported in a further systematic review, demonstrating heterogeneity in methods of existing fetal corpus callosum biometric reference [22].

Automated biometry has the potential to improve the reliability and reproducibility of derived measurements, allowing precise quantification of fetal brain development. A few recent studies have already demonstrated technical feasibility of automation of biometry for various fetal MRI measurements. The works of [23] and [24] have demonstrated automated biometry in 2-D coronal slices based on a combination of deep learning brain segmentation and machine learning morphological operations for plane selection and extraction of three linear brain measurements. In [25], atlas-based registration for propagation of landmarks was used for the extraction of 31 craniofacial biometry measurements in 3-D SVR fetal MRI head images. [26] proposed deep learning for the detection of landmarks and extraction of 11 brain measurements in 3-D SVR brain images.

However, to our knowledge, no reporting pipeline currently exists which could automate 3-D SVR T2W MRI derived biometry measurements, centile calculations and normative growth charts, and generate a clinically relevant and actionable report. Implementing reliable automated biometry directly during MRI examinations could significantly enhance clinical utility and streamline workflows, reducing the time spent acquiring manual measurements. This capability is essential for real-time integration into routine clinical practice and for maximizing the benefits of an augmented workflow.

### Contributions

This work introduces the first prototype solution for automated brain biometry and centile calculation for 3-D motion-corrected T2W fetal MRI within a clinical reporting pipeline. It includes formalization of biometry protocol based on the current clinical practice at our institution, implementation of deep learning landmark-based pipeline for computation of 13 standard biometric measurements and automated reporting of centiles based on nomograms generated from 406 control datasets across 0.55-3T field strength and 19-40 GA. The automated biometry reporting pipeline is quantitatively and qualitatively evaluated on 90 retrospective and 111 prospective cases, respectively.

## Materials and methods

### Cohort, image acquisition and preprocessing

This study utilised fetal MRI data acquired at St. Thomas’ Hospital, London across 7 ethically approved research studies: “Placental Imaging Project” (REC 16/LO/1573), “Individualised Risk prediction of adverse neonatal outcome in pregnancies that deliver preterm using advanced MRI techniques and machine learning” (REC 21/SS/0082), “CARP” (REC 19/LO/0852), “MEERKAT” (REC 21/LO/0742), “MiBirth” (REC 23/LO/0685), “NANO” (REC 22/YH/0210), and “Quantification of fetal growth and development with MRI” (REC 07/H0707/105). All studies were performed in accordance with relevant ethical guidelines and regulations. Informed written consent was obtained from all participants. For each of the datasets, T2w SST-SE/HASTE sequences were acquired on four different scanners with diverse acquisition protocols:

- 203 fetal datasets acquired on 3T Philips Achieva MRI system using a 32-channel cardiac coil with TE=180ms, acquisition resolution 1.25 x 1.25mm, slice thickness 2.5, −1.5mm gap and 5-6 stacks;
- 39 fetal datasets acquired on 1.5T Philips Ingenia MRI system using 28-channel torso coil with TE=180ms, acquisition resolution 1.25 x 1.25mm, slice thickness 2.5, −1.25mm gap, and 4-5 stacks;
- 72 fetal datasets acquired on 1.5T Siemens Sola MRI system using two 30- and 18- channel body coils with TE=80 and 180 ms, acquisition resolution 1.24 x 1.25mm, slice thickness 3mm, and 9-12 stacks;
- 212 fetal datasets acquired on 0.55T Siemens MAGNETOM Free.Max MRI system using 6-element flexible coil and a 9-element spine with TE=105–106ms, acquisition resolution 1.48 x 1.48mm, slice thickness 4.5, and 9-12 stacks [27].

The inclusion criteria for the training and testing cohorts were: (1) healthy volunteer (2) singleton pregnancy, (3) more than four fetal MRI stacks with full brain region of interest coverage, (4) acceptable SVR reconstruction quality [16] with sufficient visibility of the major brain structures and tissue interfaces and (5) normal brain appearances. Additional criteria for the prospective cohort that included a subset of cases referred for clinical indications were no reported medical conditions or neonatal complications.

The cohorts included a total of 525 MRI examinations from 18-41 weeks GA: 150 datasets were used for training of the model, 90 datasets for retrospective testing, 111 cases for prospective evaluation. 406 healthy control datasets were used to generate the normative growth charts (and included training and testing cases). All cases were de-identified prior to processing through the pipeline.

All images were 3-D reconstructed based on the fully automated auto-proc-SVRTK^1^ pipeline [28, 29] to 0.8-1.0mm resolution in the standard radiological space. In addition, for each of the image volumes we run 3-D brain tissue segmentation using “brain volumetry and automated parcellation for 3-D fetal MRI” (BOUNTI) method [30].

### Formalization of landmark-based biometry protocol

The current clinical practice at our institution involves 2D biometric measurements taken on 3D SVR images, as this approach provides full brain coverage, which is essential for reliable biometry [17].

Guided by the protocol defined in [17], we sought to expand on this framework through workshops with local reporting radiologists who have extensive experience in fetal MRI (AL, SA, AEC, MR) in order to establish a reproducible biometric protocol. For each biometric parameter, the anatomical landmarks, reference planes and slices were clearly defined across all GAs, with attention made to accounting for partial volume effects.

We incorporate additional measurements including the anteroposterior diameter of the pons, corpus callosum length, and cavum septum pellucidum width. Skull measurements were also updated to reflect outer-to-outer boundaries[31, 32]. These additional parameters were selected due to their relevance in monitoring normal fetal growth and development ([33].

Our finalized protocol includes 13 routine linear measurements of the skull, brain, ventricles, corpus callosum, cerebellum, pons and vermis (including left and right paired measurements to account for potential asymmetries).

Each measurement was defined for a subset of cases at 20-35 weeks GA using a landmark-based approach with 3-D-sphere point labels manually created in ITK-SNAP^2^. This was followed by assessment of inter- and intra-observer variability to ensure reliability and reproducibility.

### Automated brain biometry pipeline

The proposed pipeline for automated biometry for 3D T2W fetal brain images is illustrated in Fig. 2. At first, 3-D SVR-reconstructed T2W brain images are aligned to the standard radiological planes and resampled to 0.5mm resolution. Alignment is performed in MIRTK^3^ toolbox based on rigid registration of cerebral hemisphere labels extracted from brain tissue BOUNTI labels [30] to the GA-matched fetal brain atlases [34]. This is followed by automated segmentation of 26 3-D landmarks using a classical implementation of 3-D UNet [35] in MONAI[36] framework. We used the standard network architecture with four encoder-decoder blocks (output channels 16, 32, 64, 128, 256), correspondingly, convolution and upsampling kernel size of 3, ReLU activation, dropout ratio of 0.5, metric, AdamW optimiser with a linearly decaying learning rate, initialised at 1 *×* 10*^−^*^3^, default *β* parameters and weight decay=1 *×* 10*^−^*^5^, combined Dice and cross entropy loss function and standard MONAI intensity augmentations. The pre-processing includes label-based affine registration to the atlas space, masking to the brain ROI, resampling with padding to 256x256x256 grid and rescaling to 0-1 range.

**Fig. 1:**
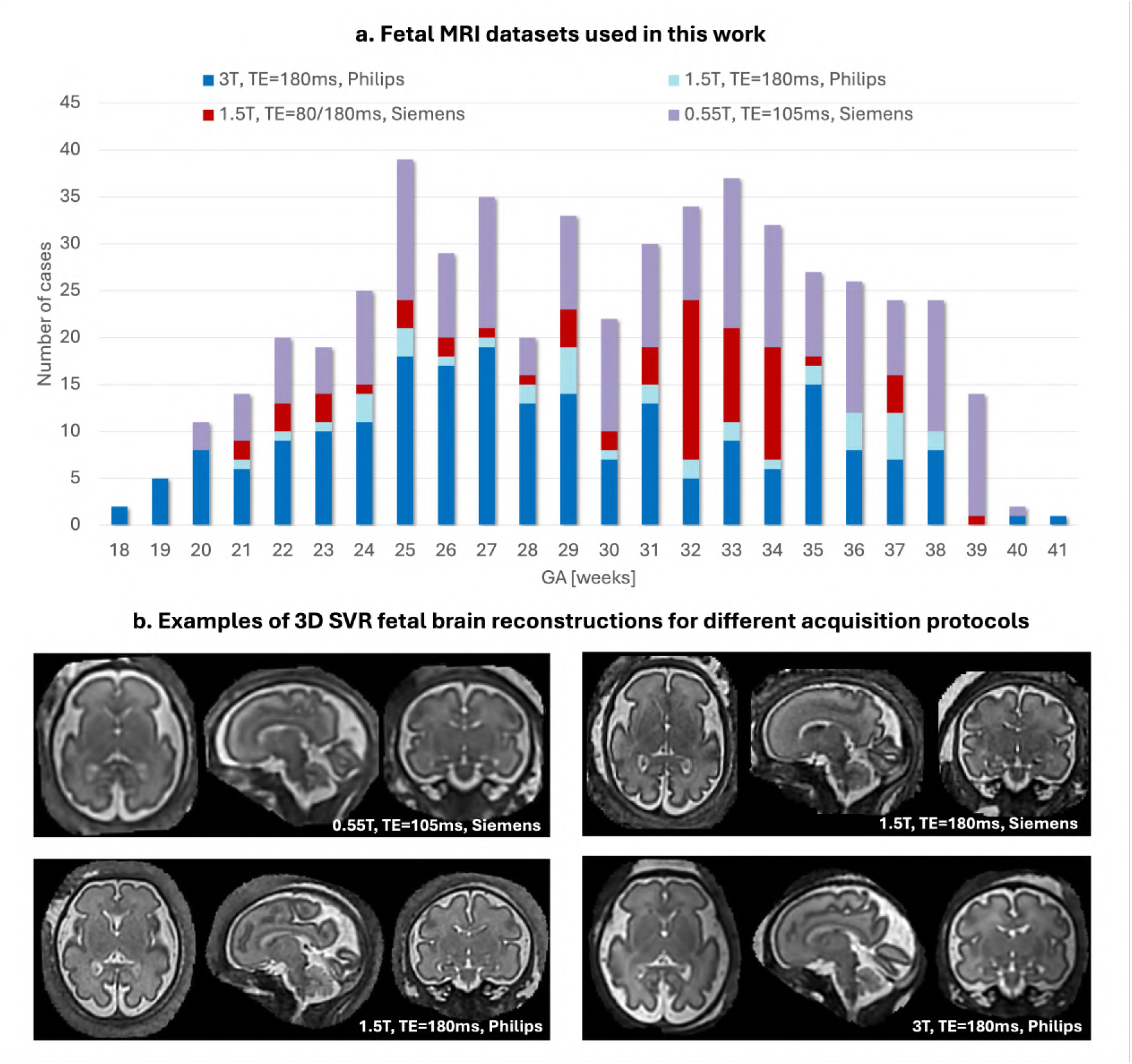
a Gestational age distribution of 525 0.55, 1.5 and 3T T2W datasets used in this work for the training of the deep learning model, generation of growth charts, retrospective quantitative testing and prospective qualitative evaluation. b Examples of 3D SVR fetal brain reconstruction for different acquisition protocols. *3-D* 3-dimensional, *GA* gestational age, *MRI* magnetic resonance imaging, *SVR* slice-to-volume registration, *T* Tesla, *TE* echo time, *T2W* T2-weighted

**Fig. 2:**
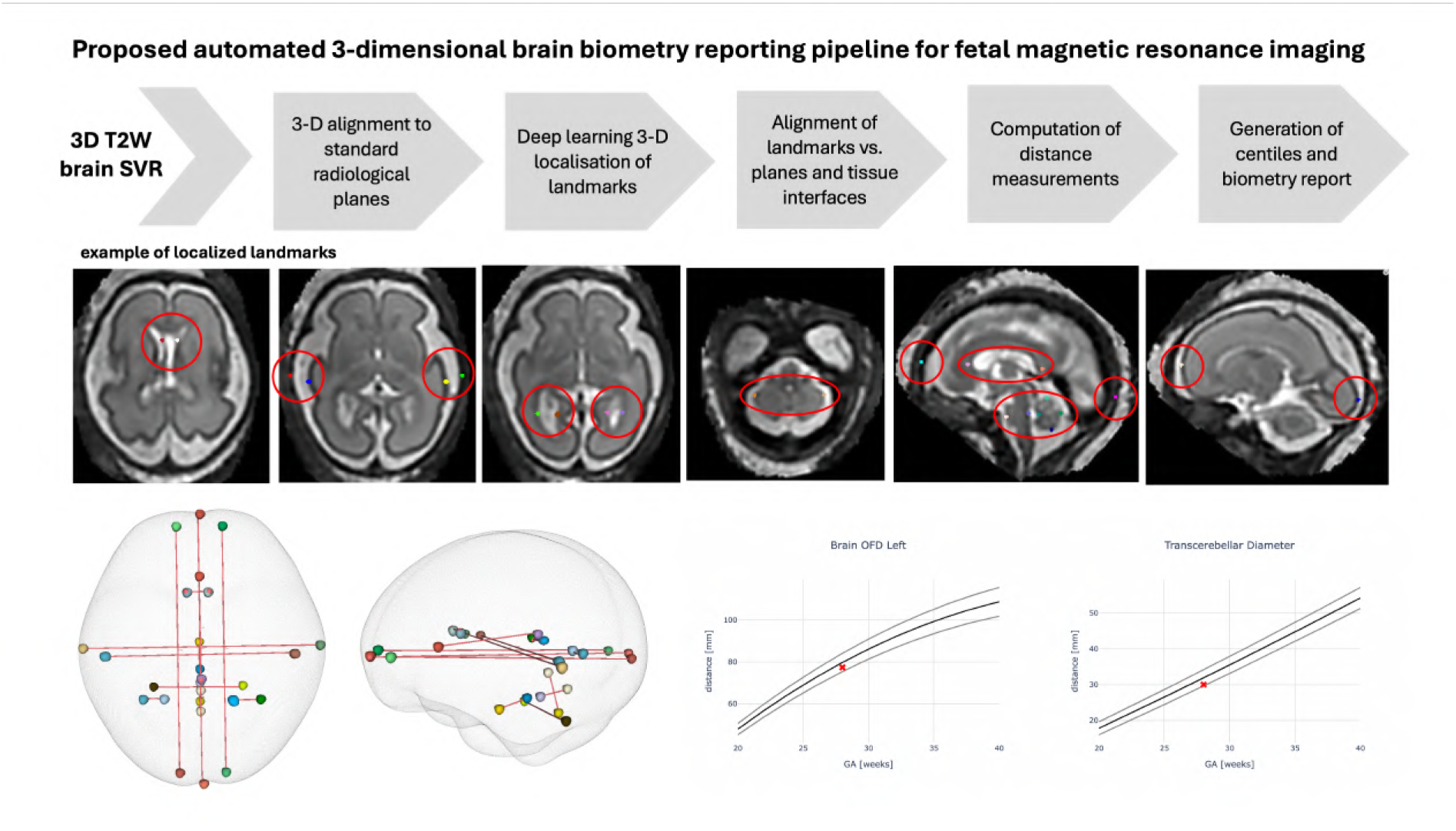
Proposed pipeline for automated brain biometry for 3-D T2W fetal MRI. *SVR* slice-to-volume reconstruction, *3-D* 3-dimensional, *T2W* T2-weighted

The network was trained on 150 0.55T and 3T datasets from 18-41 weeks GA in three stages. At first, the network was pre-trained on 30 cases with landmark labels generated by label propagation and manual refinement. This was followed by two more stages of training on datasets with manually refined previous network outputs. Next, all segmented landmarks were localised and aligned to the relevant planes and tissue interfaces based on BOUNTI brain tissue labels. The final measurements are computed in mm as distances between 3-D sphere label centre coordinates based on the biometry protocol defined above. All steps are combined in one docker-based script publicly available at *auto-proc-SVRTK* repository https://github.com/SVRTK/auto-proc-svrtk.

### Normative growth charts for fetal brain biometry

In order to provide reference ranges for the reporting tool, we used the proposed pipeline to generate normative biometry growth chart models with 5th, 50th, 95th centiles from 406 healthy control datasets (including 0.55T, 1.5T and 3T) from 18 to 40 weeks weeks GA range. All automated landmarks were reviewed and manually refined, if required. The models were generated using classical quadratic polynomial fitting [37].

### Automated biometry reporting and normative growth charts

The integration of the proposed automated biometry pipeline into routine clinical practice would require an intuitive output format, such as a report, which is easy to interpret and supports further analysis on an individual case basis, thereby enhancing the real-world applicability and acceptability.

Therefore, after the landmark detection step, the re-oriented 3-D brain image and 3-D biometry landmarks files are passed to the reporting python script that automatically generates an .html report page. The report displays:

- general information about the case (patient ID, scan date and GA);
- 8 representative 3-D brain SVR images with automated landmark-based 2D linear measurements overlaid;
- table with all extracted biometry measurements, computed centiles and z-scores;
- growth charts of all 12 extracted measurements vs. normative curve with centiles;
- disclaimer that the current report is for research purposes only, and hence should not be used as a diagnostic tool (Awaiting regulatory approval for routine clinical use).

The full pipeline combining automated biometry and reporting scripts is publicly available in a standalone central processing unit (CPU) SVRTK^4^ docker operational on Windows, MAC and Linux operational systems. The source code with instruction is publicly available online at auto-SVRTK^5^ GitHub repository.

### Evaluation

At first, the general feasibility of the implemented automated biometry pipeline was quantitatively tested on 90 retrospective cases using normal control datasets acquired with three different protocols at 0.55T, 1.5T, and 3T field strengths and covering a 22 - 38 week GA range. For all cases, two sets of manual linear measurements were performed by experienced neuroradiologists (AL, SA, AE, MR) and researcher in fetal MRI (AU). Comparison was performed between the manual measurements as well as the automated vs. the average manual measurements. The differences were computed as the absolute (mm) errors.

Next, we performed qualitative prospective evaluation of the clinical utility and acceptability of the pipeline on 111 cases acquired at St.Thomas’ Hospital, London between July and December 2024. Selection criteria included consent for research (healthy volunteer and those referred for clinical reasons), singleton pregnancy, a GA range of 20 to 38 weeks, good reconstruction quality with clear visibility of brain structures and no extreme structural abnormalities. For each of the cases, the usability of the generated automated biometry report was evaluated, and the visual inspection of all automated landmark placements was assessed using a Likert scale by 4 experienced neuroradiologists (AL, SA, AEC, MR) with 4 - 30 years in fetal and neonatal MRI and a researcher in fetal MRI (AU) with 6 years of experience. Additionally, end-user feedback was gathered to evaluate perceptions regarding the pipeline’s efficiency, its impact on workload, and overall confidence levels.

Manual measurements and review of landmark placements were performed in ITK-SNAP, 3D Slicer^6^ and MITK Workbench^7^ platforms for medical image visualization.

## Results

### Landmark-based fetal brain biometry protocol

Fig. 3-4 present the proposed standardized protocol with definitions of 13 routine fetal measurements (including left and right atrial diameter and brain occipito-frontal distance) and representative images at 20 - 35 week time points.

**Fig. 3:**
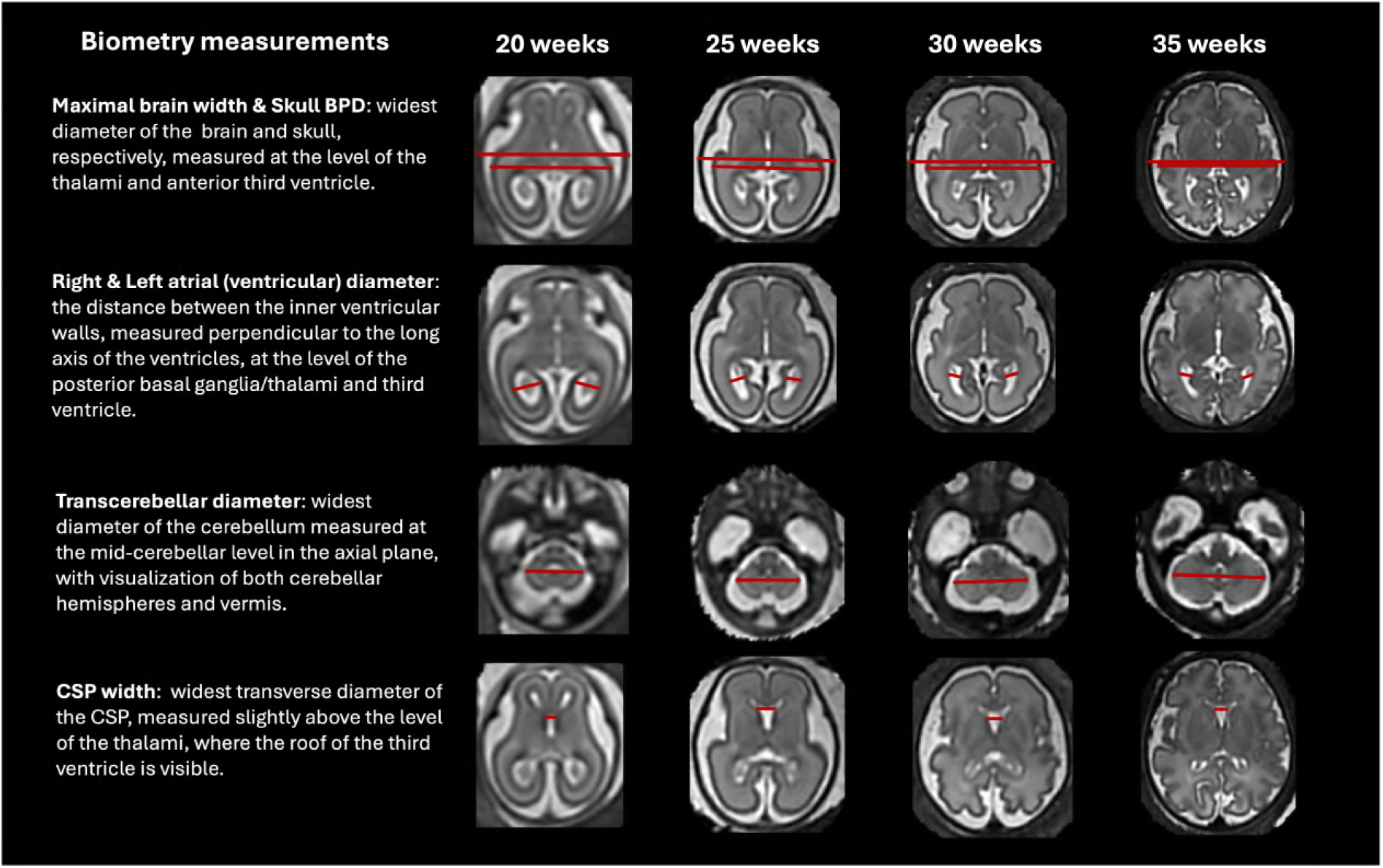
Proposed formalized protocol for automated brain biometry for 3-D T2W fetal MRI from 20 to 35 weeks GA range (axial planes, part 1). *BPD* bi-parietal diameter, *CSP* cavum septum pellucidum

**Fig. 4:**
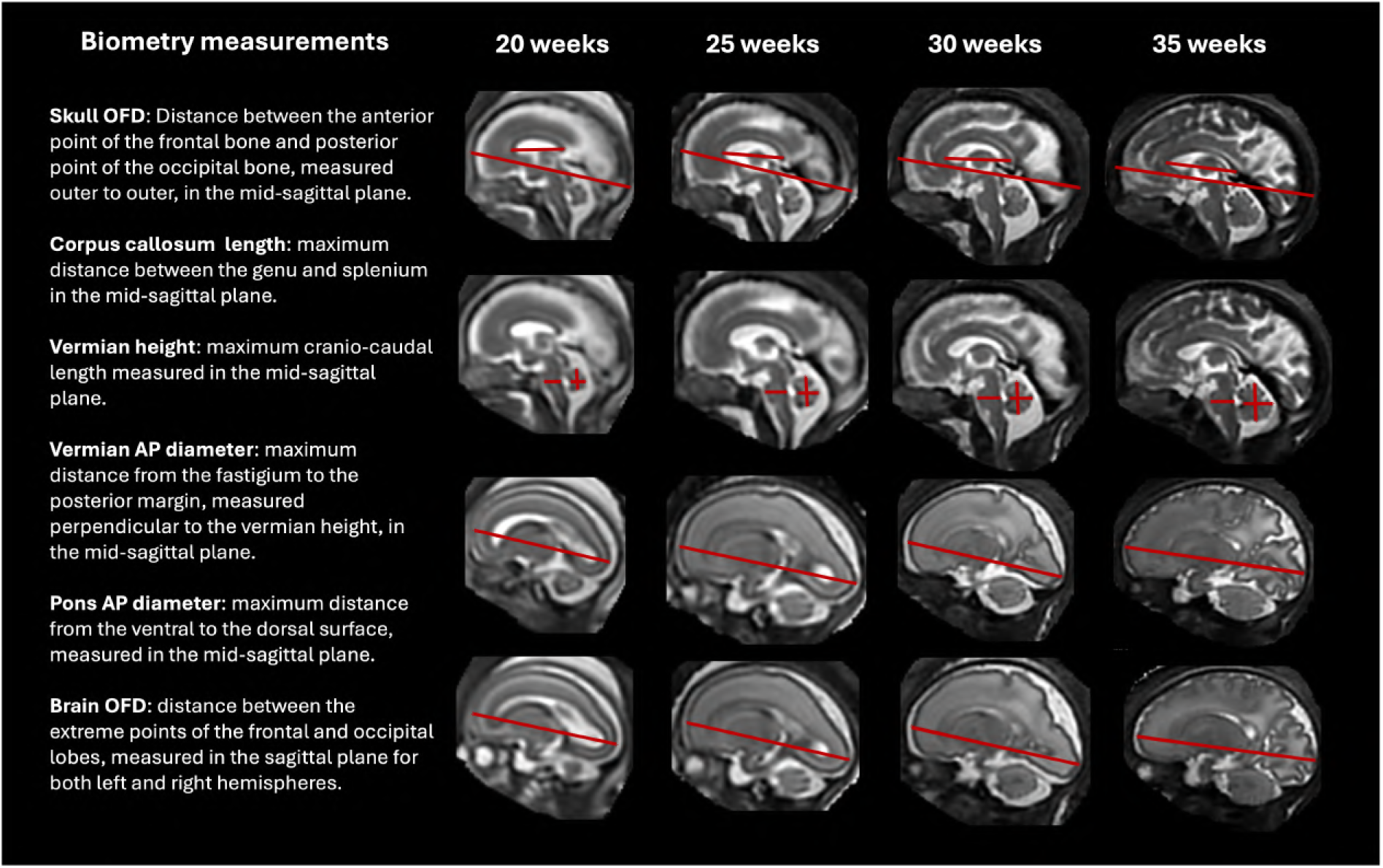
Proposed formalised protocol for automated brain biometry for 3-D T2W fetal MRI from 20 to 35 weeks GA range (sagittal planes, part 2). *AP* antero-posterior, *OFD* occipito-frontal diameter

The biometric parameters were chosen for their relevance in monitoring normal fetal growth and development, aiding in the diagnosis of common fetal abnormalities such as ventriculomegaly, corpus callosum agenesis and posterior fossa malformations as well as in prognosis prediction. It builds on the work outlined by [17], updating measurement definitions to reflect the current standard clinical practice at our institution, including renaming brain biparietal diameter to maximal brain width. Additionally, key parameters routinely measured were incorporated. The updated definitions also account for the dynamic morphology with gestation, partial volume effects, and variations in field strength.

- The skull biparietal diameter (BPD) is defined as the widest diameter of the skull and is measured in the axial plane at the level of the thalami and anterior third ventricle, from outer to outer. The skull occipito-frontal diameter (OFD) is defined as the distance between the anterior point of the frontal bone and the posterior point of the occipital bone, measured in the mid-sagittal plane from outer to outer.
- The maximal brain width is the widest diameter of the brain, measured at the level of the thalami and anterior third ventricle in the axial plane. The brain OFD is the distance between the extreme points of the frontal and occipital lobes, measured separately for the left and right hemispheres in the sagittal plane.
- The right and left atrial (ventricular) diameters are measured as the distance between the inner ventricular walls, perpendicular to the long axis of the ventricles, at the level of the posterior basal ganglia, thalami, and third ventricle. The cavum septum pellucidum (CSP) width is defined as the widest transverse diameter of the CSP, between the inner margins of the septal leaflets. It is measured slightly above the level of the thalami, where the roof of the third ventricle is visible.
- The transcerebellar diameter (TCD) is defined as the widest diameter of the cerebellum and is measured at the mid-cerebellar level in the axial plane, ensuring visualization of both cerebellar hemispheres and the vermis. The vermian height is the maximum cranio-caudal length, and the vermian antero-posterior (AP) diameter is the maximum distance from the fastigium to the posterior margin, measured perpendicular to the vermian height, both obtained in the mid-sagittal plane.
- The pons antero-posterior diameter is the maximum distance from the ventral to the dorsal surface, measured in the mid-sagittal plane. The corpus callosum length is the maximum distance between the genu and splenium, also measured in the mid-sagittal plane.

### Retrospective evaluation of the automated biometry pipeline

Fig. 5 summarizes the results of testing of the pipeline on 90 retrospective from 0.55T, 1.5T and 3T protocol and 22-38 weeks GA range for automated vs. two sets of manual measurements. The intra-class correlation coefficient (ICC) values showed good to excellent reliability for most of the measurements and are comparable between both manual vs. manual and automated vs. average manual measurements for all 13 biometrics. The distribution of the absolute error was also similar between manual with presence of several outliers. The range of maximum absolute errors compared to manual measurements ranged from 1 to 3 mm, consistent with the previously reported variability in automated methods [23, 24, 26]. Examples of localized landmarks for test cases at different acquisition protocols and GAs are shown in Fig. 6. The placement of the automated landmarks in all cases was rated (AL) as good or acceptable. The measurements with the highest degree of uncertainty were the maximal brain width, ventricular (atrial) diameters and cavum width due to the anatomical variability and the absence of clear fixed structural landmark points in 3D even after formalisation of the protocol. Atrial diameters and cavum width also have lower ICC for the interobserver variability. This indicates that while these results confirm general feasibility of the automated biometry vs. manual measurements, further work is required for establishing of a biometry protocol specifically adapted for 3D SVR images. Moreover,

**Fig. 5:**
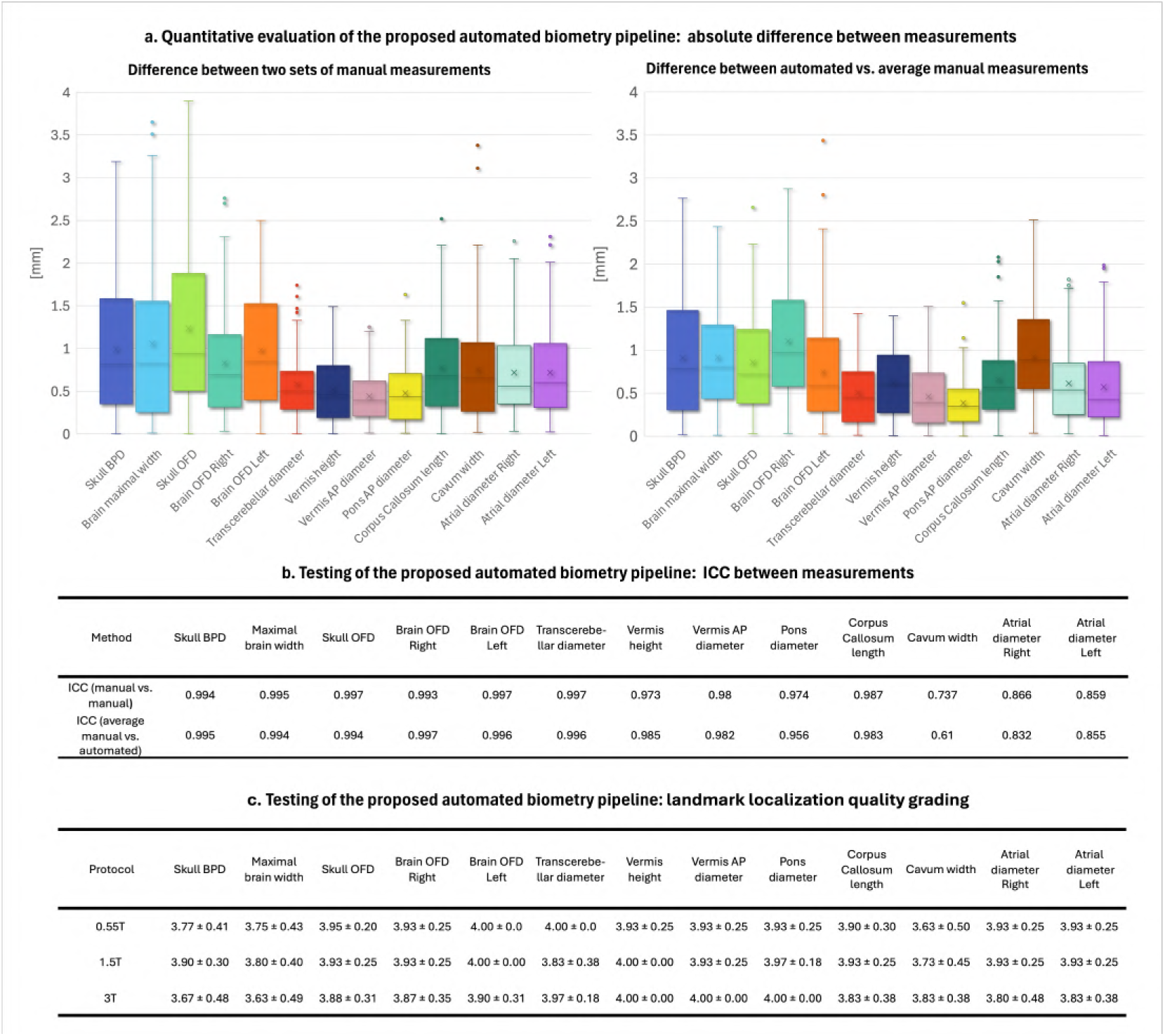
Testing of the proposed automated biometry pipeline on 90 0.55T/1.5T/3T cases from 22 to 38 weeks GA. a Absolute difference between two sets of manuals measurements and absolute difference between average manual vs. automated measurements. b ICC values between two sets of manuals measurements and between average manual vs. automated measurements c Landmark localisation quality grading (*four* good; *three* acceptable; *two* poor; *one* fail). *AP* antero-posterior, *T* tesla, *GA* gestational age

**Fig. 6:**
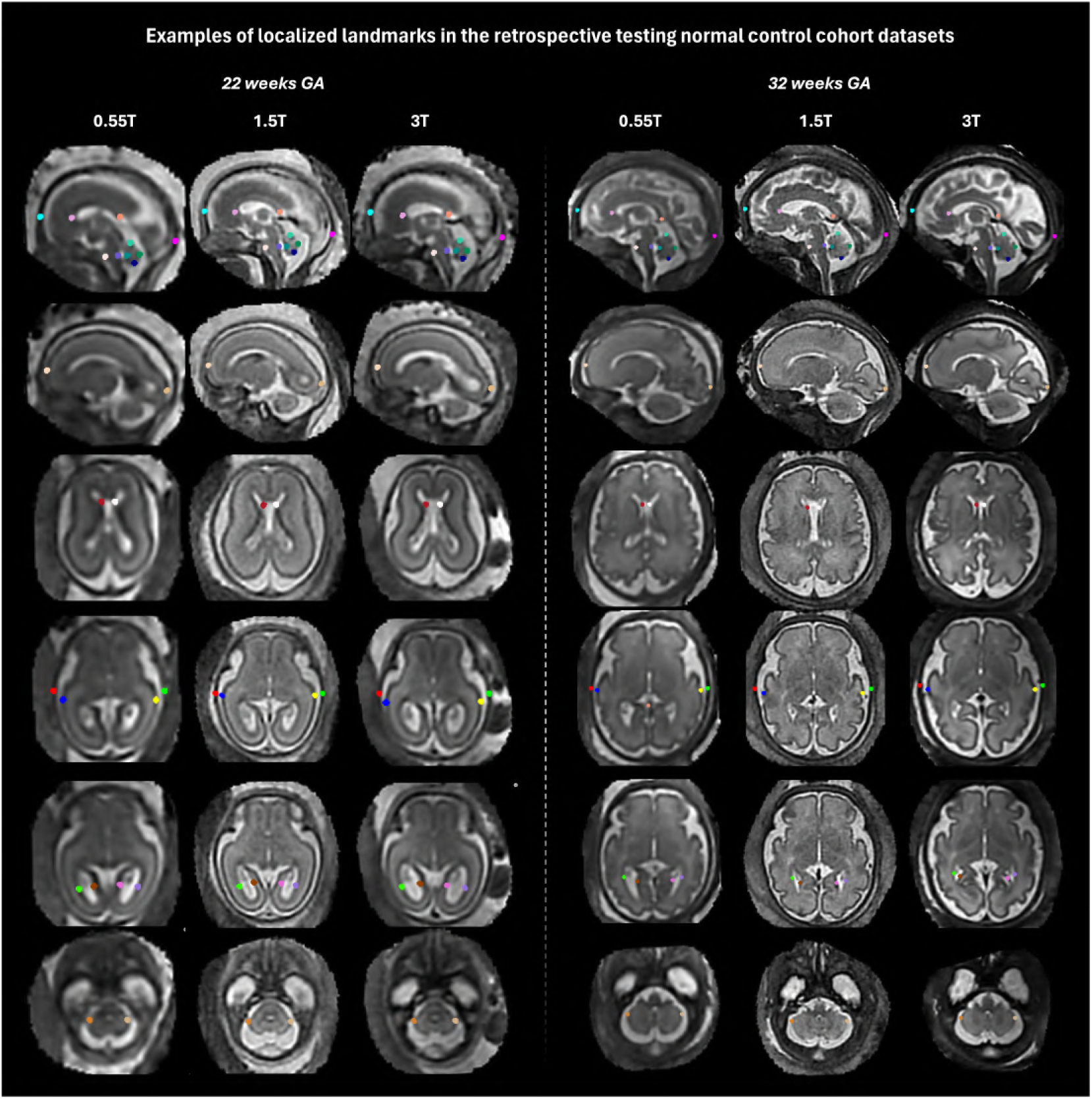
Testing of the proposed automated biometry pipeline on 90 0.55T/1.5T/3T cases from 22 to 38 weeks GA: examples of localized landmarks in different acquisition protocols and GAs. *T* tesla, *GA* gestational age further optimisation of the model for anatomical variations along with the additional automated quality control for image quality and landmark placement certainty grading will be required to achieve stable performance.

### Normative growth charts for fetal brain biometry

Fig. 7-8 displays the generated growth charts and quadratic normative models created from automated biometry measurements from 406 normal control datasets from 18 to 40 weeks GA range. All biometry results were inspected and minor manual editing of the landmarks was required in 53 cases due to suboptimal image quality and low visibility, which did not result in significant difference on the trendlines. The numerical scales and shape characteristics of the models highly correlate with the trends previously reported in Kyriakopoulou et al. [17] and have similar appearance to the trends in the earlier guidelines [10–13, 15].

**Fig. 7:**
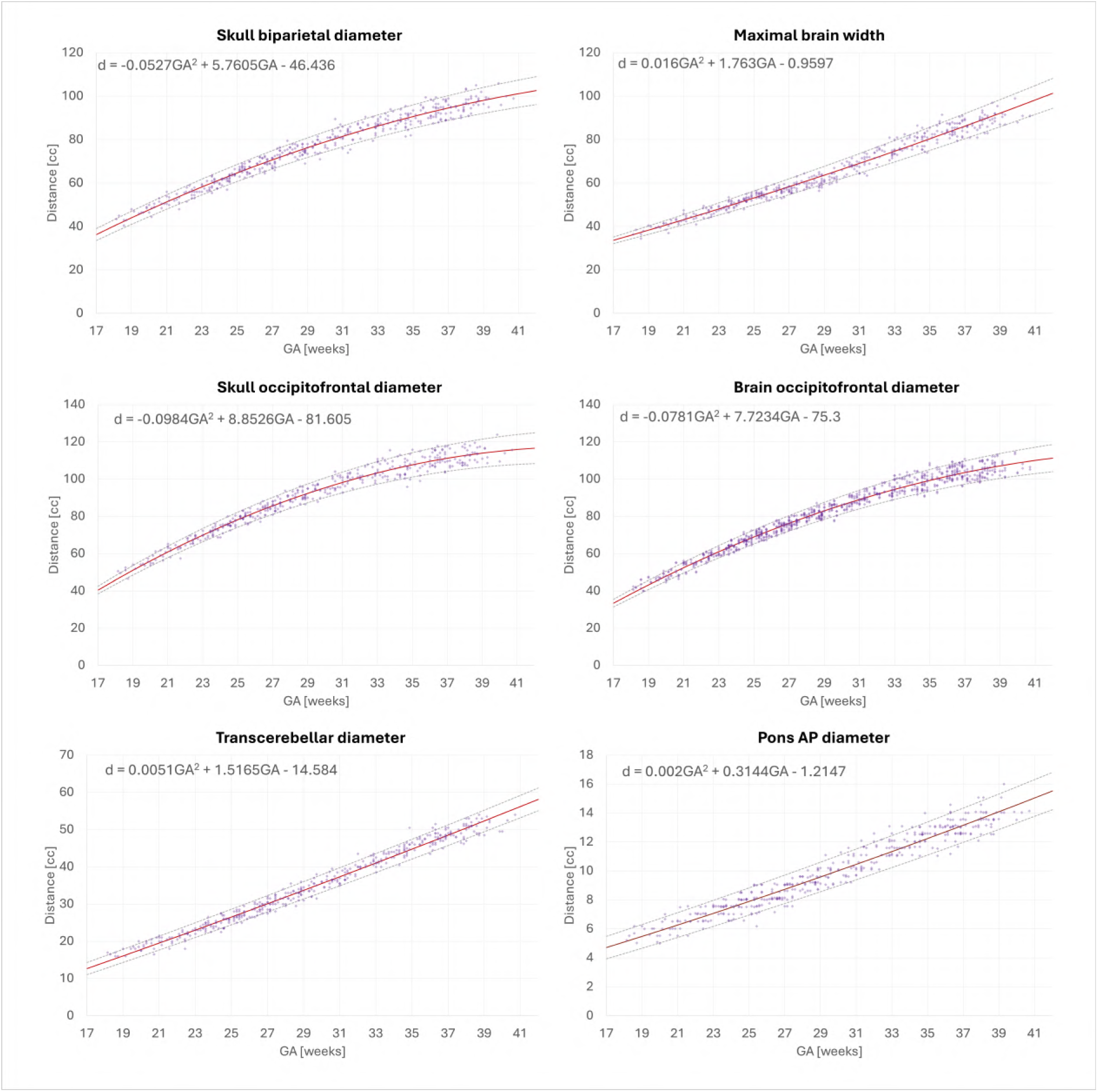
Normative growth charts created from automated biometry for 406 control (0.55, 1.5 and 3T) subjects with centiles (part 1). *AP* antero-posterior, *GA* gestational age

**Fig. 8:**
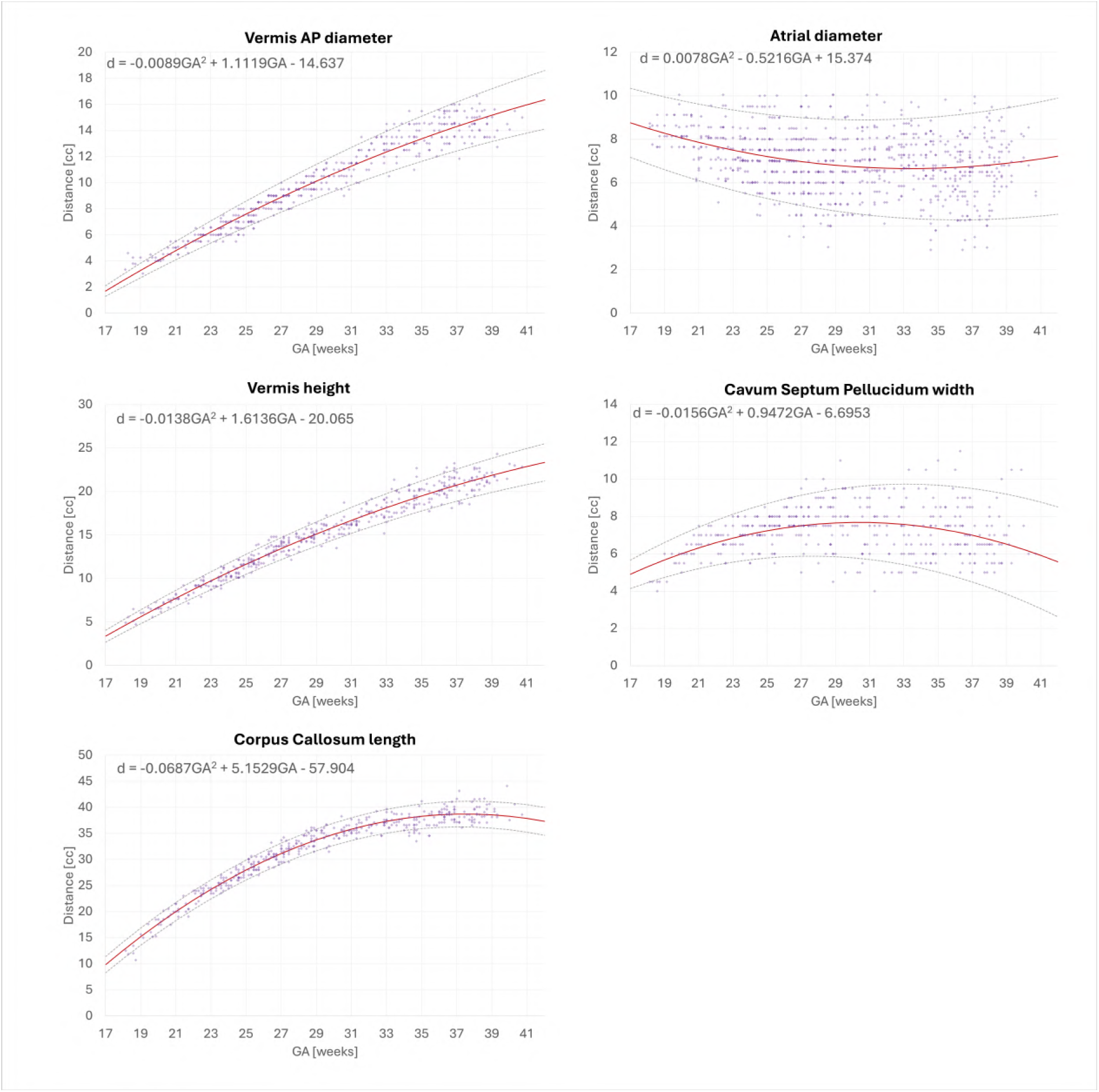
Normative growth charts created from automated biometry for 406 control (0.55, 1.5 and 3T) subjects with centiles (part 1). *AP* antero-posterior, *GA* gestational age

All global measurements including skull BPD, skull OFD, brain OFD and brain maximal width constantly increase with GA with growth trajectory slightly decelerating towards late gestation. The transcerebellar diameter, pons width, vermian width and vermian height trends also show almost linear growth in size. The corpus callosum length trajectory tends to plateau after approximately 33 weeks. The ventricle (atrial) diameter values show high variance during the whole GA range also reported in previous works [17]. The global trend demonstrates that ventricles are larger in earlier GA and tendency to reduce over gestation. There is also a high variance in cavum width which plateaus after the third trimester.

The document with centile model formulas based on the mixed acquisition protocol datasets together with examples of landmark biometry protocol defined in the atlas space are publicly available online at a dedicated repository https://gin.g-node.org/ kcl cdb/fetal mri biometry.

### Prospective evaluation of the biometry reporting pipeline

We successfully ran automated biometry reporting pipeline for 111 prospective datasets without reported extreme structural brain anomalies. Qualitative grades and comments were collected from the evaluators about the landmark placement (26 in total) and general end-user feedback on utility and practicality of the reporting pipeline.

The resulting average quality scores for each of the 26 landmarks of 13 measurements are presented in Fig. 9. On average, more than 98% of all landmark placement were graded as acceptable for interpretation and measurements (65.9% “excellent”, 26.1% “good”, 6.8% “fair” grades). Only 1.1% of landmarks were graded as “poor”. The best performance with the highest proportion of “excellent” grades was recorded for skull OFD and BPD measurements, TCD, pons and vermis AP diameters and vermis height. This can be attributed to the pronounced and clearly visible anatomical features that are required for accurate placement of landmarks. In comparison, the maximal brain width and brain OFD landmarks expectedly had higher proportion of “good” grades due to inherently higher morphological variations and increased cortical folding that occur with gestation. The lowest summary grades, but still graded as “good”, were observed for the cavum and atrial diameter measurements due to anatomical variability, and corpus callosum due to suboptimal visibility of the genu.

**Fig. 9:**
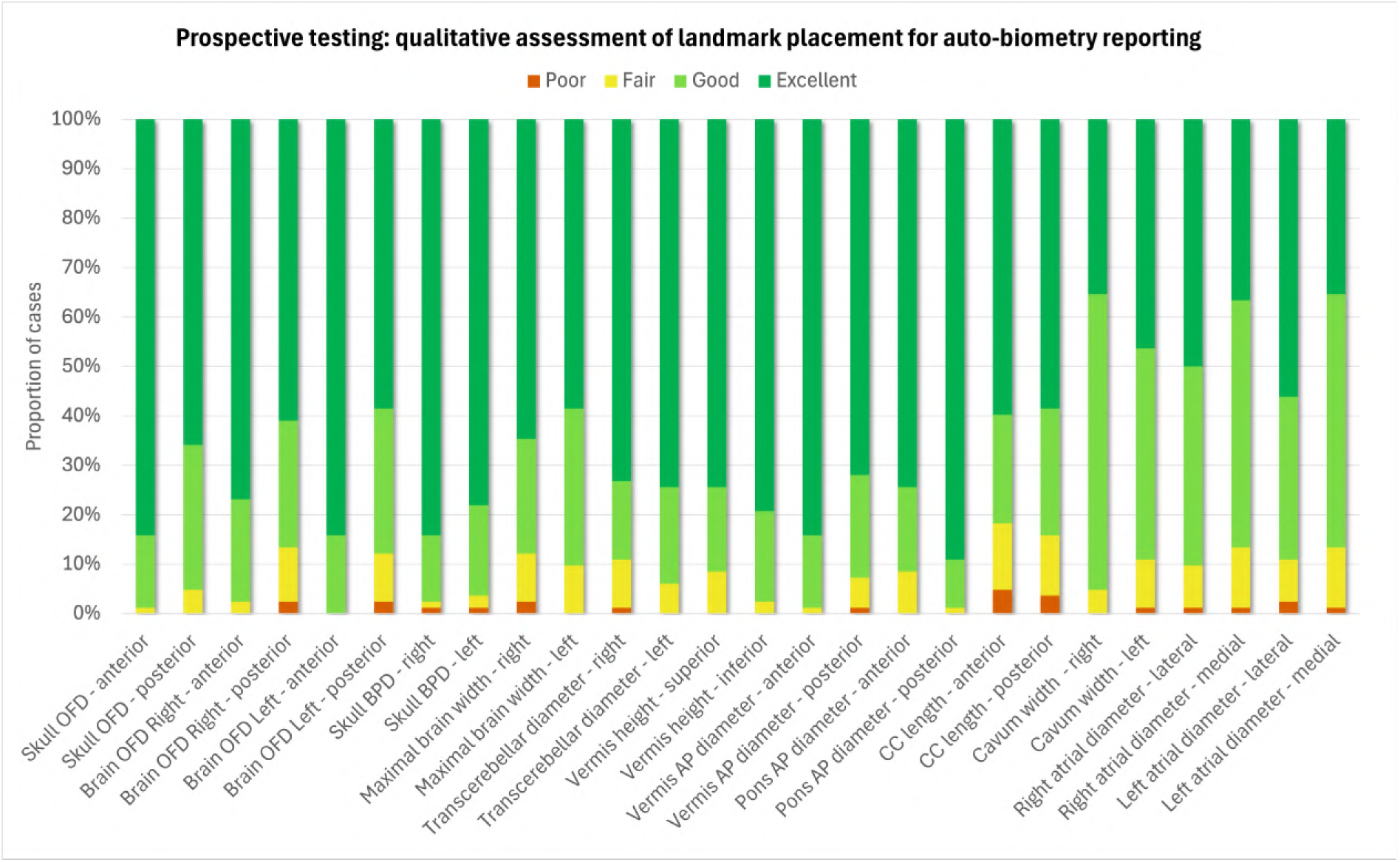
Results of qualitative evaluation of landmark placement for 111 prospective datasets from 0.55T, 1.5T and 3T acquisition protocols. Quality grades: poor, fair, good, excellent. *AP* antero-posterior, *BPD* bi-parietal diameter, *CC* corpus callosum, *OFD* occipito-frontal diameter

These results reaffirm that using automated biometry reporting in clinical practice is feasible and that the proposed pipeline can be potentially used as a baseline for further adaptation and retraining of the model for a wide range of brain anomalies. Yet, the minor proportion of low quality grades highlights that automated measurements should always be visually inspected and corrected, if required, prior to reporting.

An example of the generated automated biometry .html report for one of the cases is shown in Fig. 10. The format was confirmed as generally acceptable for clinical interpretation. The total processing time for the full pipeline including preprocessing, deep learning biometry and report generation is less than five minutes per case on average which is within reasonable ranges for reporting.

**Fig. 10:**
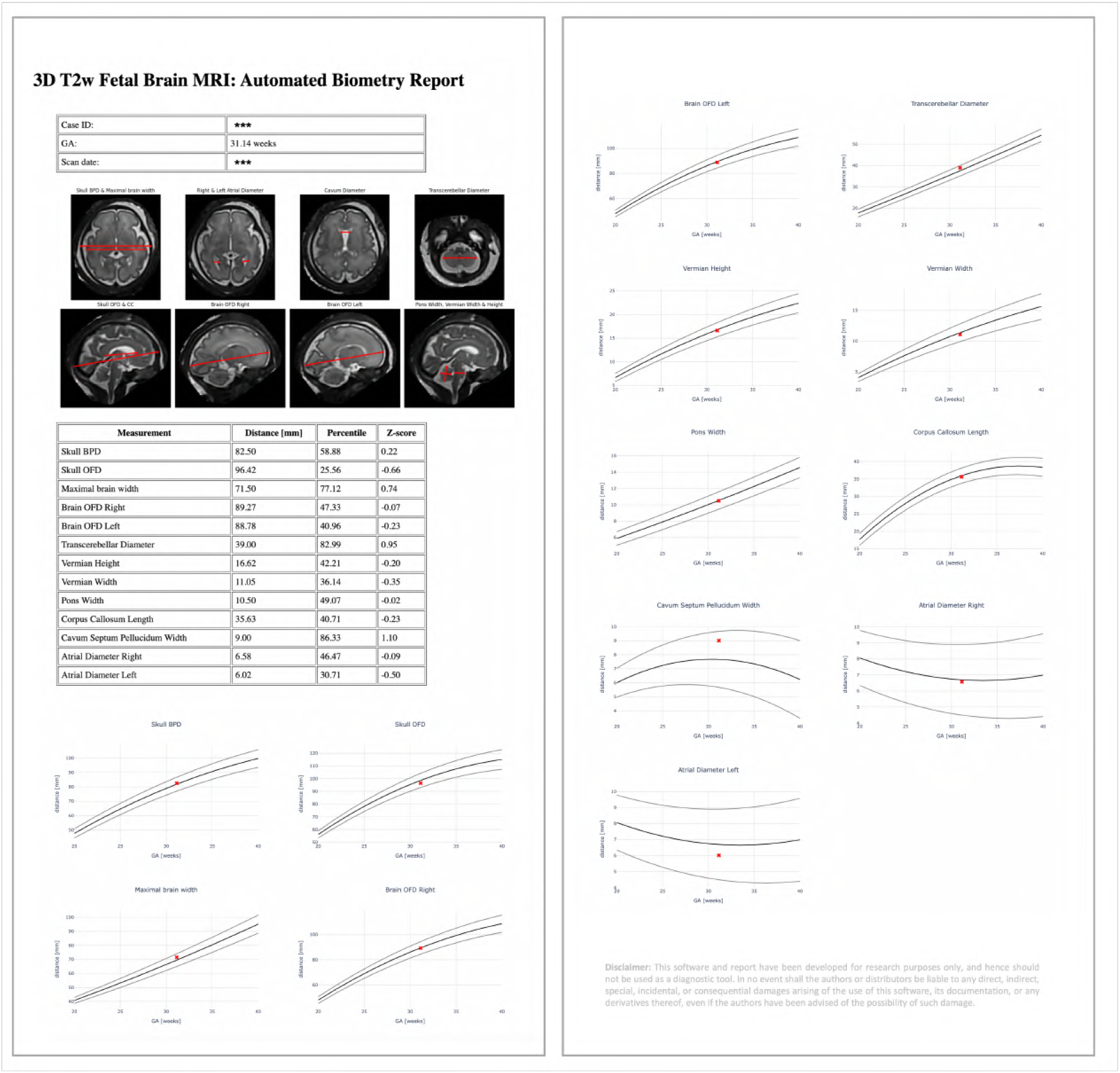
Example of the automated report for a test case generated as a .html file. *BPD* bi-parietal diameter, *GA* gestational age, *OFD* occipito-frontal diameter

## Discussion

This work introduced the first prototype pipeline for a fully automated, end-to-end, brain biometry and centile calculation for 3-D motion-corrected T2W fetal MRI across different field strengths and gestational age range, within a clinical reporting pipeline. We first established the landmark-based protocol with 13 routine linear measurements (including paired measurements) through expert consensus with experienced fetal neuroradiologists. Each measurement was clearly defined by determining precise and reproducible anatomical landmarks on reference planes and slices, and considering gestational age-related changes in morphology of the structures being measured. Next, we developed a deep learning pipeline to segment the selected landmarks for each biometric parameter and automate the measurements, which was trained on 150 cases from different acquisition protocols and a wide GA range. We implemented a script for automated reporting that displays the biometry results and centile calculation, with the output presented in .html format, available to the end-user at the time of reporting as a decision support tool to help guide accurate consistent and biometry assessment. The performance of the pipeline was evaluated on a retrospective test cohort of 90 cases, while its clinical utility and acceptability assessed on a prospective cohort of 111 cases.

Our results demonstrate good accuracy, with the range of absolute errors compared to manual measurements varying from 1 to 3 mm, consistent with the previously reported variability in automated fetal brain biometry methods. The prospective evaluation assessing the placement of each landmark was also considered good or excellent for more than 90% of all landmarks in all datasets. In both retrospective testing and prospective evaluation, the measurements with the highest degree of variability were the maximal brain width, corpus callosum length, ventricular atrial diameters and cavum width. While we defined the landmarks across all measurements, the absence of fixed and clearly visible anatomical structures for the above parameters may have introduced inconsistencies in model identification and subsequently reduced reproducibility. Other factors contributing to variability include partial volume effects, image quality and normal anatomical variability between fetuses. However, for the smaller brain structures, such as the vermis and pons, the results were favourable, with consistent performance and quality assessments rated as good or excellent. These structures have traditionally posed challenges for accurate assessments on 2-D slices due to the difficulty in obtaining true orthogonal mid-sagittal planes. The ability to reliably assess such measurements demonstrates the potential for improved biometry accuracy using automation with SVR reconstructions.

Furthermore, the average time to complete all manual measurements is 10-15 minutes for an experienced fetal neuroradiologist, while inspection of the automated landmark placement and computed biometrics took only 1-3 minutes per case. Therefore, even in cases where certain parameters may require re-measurement, the overall time savings and efficiency gains are substantial.

The main strengths of this work include a large normative dataset which spans a wide GA range, encompasses diverse acquisition parameters and incorporates 3-D SVR. The utilisation of 3-D SVR reconstruction has also shown to improve the accuracy of fetal brain biometry by addressing the technical limitations of traditional 2-D slice based measurements [17, 18].

The original methodology for biometry measurements for fetal brain MRI [10] was proposed more than 20 years ago on 2-D slices, developed in accordance to ultrasound practices. This methodology was also derived from fetuses referred for suspected abnormalities, on single field strength with limited sample sizes at extreme gestational ages. Accurate assessments on 2D images rely on the acquisition of true orthogonal planes, which are often challenging due to fetal and maternal motion. Furthermore, the optimal slice may not always be captured as a result of inter-slice motion, resulting in underestimation of the measurement.

A further limitation of this methodology is that it does not address the dynamic changes in fetal brain morphology, making it difficult to apply consistent reference points across different GA. For example, Garel ([10] defines the measurement of the lateral ventricles as being taken at the level of the atria (with good visibility of the choroid plexuses), on an axis perpendicular to that of the ventricle and at the midheight of the ventricle. However, at younger GAs, the orientation of the ventricles is more vertical making it difficult to use the same axis defined for later gestation when the shape adopts the adult-like orientation.

By refining and expanding on the biometry protocol proposed by [17], our work addressed this challenge by accounting for the developmental changes in brain structures, demonstrating reproducible automated biometry across different GAs.

Moreover, given the vast amount structural information that can now be extracted from fetal MRI scans, establishing MRI-specific normative reference ranges would not only improve the accuracy and reliability of biometric measurements, but also capture the complex and dynamic process of fetal brain development. The generated normative growth charts based on the proposed automated biometry pipeline from 406 control subjects from 18 - 40 weeks GA range demonstrate high correlation with the trends in the previously reported works.

Compared to previous published work on automated biometry, which narrowly focus on technical performance, we have developed an enhanced solution by implementing a comprehensive end-to-end pipeline. We have also demonstrated the feasibility of this pipeline in a prospective clinical setting, facilitating real-time integration into clinical workflows and streamlining workflows, offering operational benefits such as improved efficiency.

Our work represents a key step towards the standardization of automated fetal brain biometry reporting, addressing a critical gap in current clinical practice by reducing reliance on manual approaches and ensuring accurate, reliable and reproducible measurements to enhance the full clinical report. Furthermore, the end-to-end pipeline can overcome the practical challenges to widespread clinical integration, which would enable more centres to undertake good quality fetal MRI, as well as increasing accessibility to a broader patient population.

### Limitations and future work

While we have demonstrated the initial feasibility of an automated biometry pipeline for fetal brain MRI, it represents only a part of the full clincial report and in addition extensive further work is needed before it is ready for routine integration into clinical practice. This includes incorporating abnormal cases with structural anomalies and cases with varying imaging quality to ensure robustness, as well as validating the pipeline on external datasets with diverse patient populations to improve generalizability. Furthermore, expanding the scope to include craniofacial biometry [25] as well as volumetric information and 3-D surface-based biometry measurements based on segmentations [30] could further enhance its clinical utility, enabling more precise quantification of brain development as well as improved detection and characterization of abnormalities. In addition, an automated quality control system would also need to be implemented to detect pipeline issues, failure modes and model performance drift. We also will need to correlate with neurodevelopmental outcomes, and explore sex, parental characteristics, ethnicity differences and other determinants which can impact normal fetal growth. These variables are critical for assessing the robustness of the automated system, ensuring it is applicable across diverse populations.

## Conclusion

This work introduces the first prototype for automated brain biometry and centile calculation for 3-D motion-corrected T2w fetal MRI within a clinical reporting pipeline. The quantitative and qualitative evaluation on 90 and 111 datasets from different acquisition protocols and wide GA range confirmed the general feasibility and utility of the pipeline as a decision support tool for clinical interpretation. We also generated normative growth charts for 19 - 40 weeks GA from 406 control datasets.Our future work will focus on optimisation for various fetal brain anomalies, extension of the list of measurements, development of automated quality control and external validation of the reporting tool.

## Acknowledgements

We thank everyone who was involved in acquisition and analysis of the datasets at the Department of Perinatal Imaging and Health at Kings College London and St Thomas’ Hospital. We thank all participants and their families.

This work was supported by the MRC grant [MR/X010007/1], the NIHR Advanced Fellowship to L.S. [NIHR3016640], the Wellcome Trust, Sir Henry Wellcome Fellowship to J.H. [201374/Z/16/Z], the UKRI FLF to J.H. [MR/T018119/1], DFG Heisenberg [502024488] the High Tech Agenda Bavaria to J.H., the MRC grant [MR/W019469/1], the Wellcome/EPSRC Centre [WT203148/Z/16/Z], the NIHR Clinical Research Facility (CRF) at Guy’s and St Thomas’ and by the National Institute for Health Research Biomedical Research Centre based at Guy’s and St Thomas’ NHS Foundation Trust and King’s College London.

The views expressed are those of the authors and not necessarily those of the NHS, the NIHR or the Department of Health.

## Data availability

The individual fetal MRI datasets used for this study are not publicly available due to ethics regulations. For more information please contact Jana Hutter.

## Author contributions

AL and AU contributed equally to this work. AL formalised the fetal brain biometry protocol, worked on optimisation of deep learning biometry pipeline and reporting script, performed prospective testing and qualitative evaluation and prepared the manuscript. AU developed deep learning biometry pipeline and reporting script, worked on testing and evaluation and prepared the manuscript. JM contributed to formalisation of the protocol, development of the biometry pipeline, evaluation of the reporting pipeline and prepared the manuscript. SA and AEC contributed to formalisation of the biometry protocol and evaluation of the biometry reporting pipeline. VK contributed to formalisation of the biometry protocol. SNS, JAV, MH, SM, KC participated in acquisition of the datasets. JVH supervised various parts of the project. JH and LS provided fetal MRI datasets and supervised various parts of the project.

MR provided fetal MRI datasets, contributed to formalisation of the biometry protocol and evaluation and supervised all stages of the project. All authors reviewed the manuscript.

## Declarations

The authors have no competing interests to declare that are relevant to the content of this article.

## Footnotes

1 auto-proc-SVRTK toolbox: https://github.com/SVRTK/auto-proc-svrtk

2 ITK-SNAP tool: http://www.itksnap.org/

3 MIRTK toolbox: https://github.com/BioMedIA/MIRTK

4 SVRTK docker: https://hub.docker.com/r/fetalsvrtk/svrtk

5 auto-SVRTK toolbox: https://github.com/SVRTK/auto-proc-svrtk

6 3D Slicer platform: https://www.slicer.org/

7 MITK Workbench platform: https://docs.mitk.org/2024.06/MITKWorkbenchManualPage.html

## Notes

### Competing Interest Statement

The authors have declared no competing interest.

### Author Declarations

The following relevant datasets were given research ethics committee (REC) approval by the Health Research Authority: "Placental Imaging Project" (REC 16/LO/1573), "Individualised Risk prediction of adverse neonatal outcome in pregnancies that deliver preterm using advanced MRI techniques and machine learning" (REC 21/SS/0082), "CARP" (REC 19/LO/0852), "MEERKAT" (REC 21/LO/0742), "MiBirth" (REC 23/LO/0685), "NANO" (REC 22/YH/0210), and "Quantification of fetal growth and development with MRI" (REC 07/H0707/105). All studies were performed in accordance with relevant ethical guidelines and regulations. Informed written consent was obtained from all participants.

